# Bevacizumab in ovarian cancer: time-dependent changes in risk of progression

**DOI:** 10.1101/2023.02.14.23285833

**Authors:** Shiro Takamatsu, Hidekatsu Nakai, Ken Yamaguchi, Junzo Hamanishi, Masaki Mandai, Noriomi Matsumura

## Abstract

**Background:** Bevacizumab has been used in the first-line and recurrent treatment of ovarian cancer. However, the time-dependent changes in the effects of bevacizumab administration are not fully understood.

**Methods:** The ICON7-A cohort was generated from two ancillary analyses of the ICON7 trial. ICON7-A tumors were classified as homologous recombination deficient (HRD) or non-HRD based on their gene expression profiles. Kaplan-Meier curves from published phase III trials were graphically analyzed to examine changes in the risk of progression over time between groups.

**Results:** In the ICON7-A cohort, the risk of progression in the bevacizumab group (Bev+) compared to the control group (Bev-) was lowest at 6 months. Thereafter, the risk in Bev+ gradually increased and became higher than in Bev-after discontinuation of bevacizumab at 12 months, showing a “rebound effect”. Restricted mean survival analysis showed that Bev+ had significantly better progression-free survival (PFS) than Bev-before bevacizumab discontinuation, but had significantly worse PFS after bevacizumab discontinuation. The rebound effect was observed both in HRD and non-HRD tumors of the serous subtype, but not in the non-serous subtype. In Kaplan-Meier curve image-based analysis, time-dependent changes in the risk of progression, including rebound effects, were replicated in the overall ICON7 and GOG-0218 cohorts and in their subgroups stratified by prognostic factors, presence of HR-associated mutations and chemotherapy sensitivity. In contrast, no rebound effect was observed in the studies GOG-0213, OCEANS, AURERIA and MITO16B, in which relapsed patients received bevacizumab until progression.

**Conclusion:** In ovarian cancer, bevacizumab reduces progression for approximately one year after initiation, but discontinuation may increase subsequent progression in the serous subtype regardless of HRD status. The results suggest that in the first-line treatment of ovarian cancer, bevacizumab is more beneficial in patients with a short expected survival who are unlikely to be affected by the rebound effect.

## Introduction

Ovarian cancer has the worst prognosis among gynecological malignancies ^1^. Most cases are diagnosed at an advanced stage with peritoneal dissemination and require a combination of surgery and drug therapy. The standard first-line chemotherapy is a combination of paclitaxel and carboplatin, but in the past decade, bevacizumab, an anti-VEGF-A antibody, has been incorporated with chemotherapy and used for subsequent maintenance therapy ^2^. On the other hand, high-grade serous carcinomas, which comprise the majority of ovarian cancers, are frequently associated with DNA homologous recombination deficiency (HRD), and since HRD is related to sensitivity to platinum and poly (ADP-ribose) polymerase (PARP) inhibitors ^3, 4, 5, 6^ treatment individualization based on HRD status has recently been proposed in clinical practice ^7^. Therefore, it has become more important to investigate in detail the association between therapeutic effect of bevacizumab and HRD status in ovarian cancer.

In the ICON7 trial, bevacizumab was administered at 7.5 mg/m^2^ for a total of 18 cycles (12 months) and reduced the risk of progression with a hazard ratio (HR) of 0.81 (95% confidence interval (CI); 0.70–0.94) ^8^. In the GOG-0218 study, bevacizumab was administered at 15 mg/m^2^ for a total of 22 cycles (15 months), reducing the risk of progression with HR of 0.72 (95%CI; 0.63–0.82) ^9^. However, it should be noted that since the difference in progression risk between the comparison arms significantly varied over time, the proportional hazards assumption was known to be inappropriate ^8^. Nevertheless, previous reports examining the effect of bevacizumab, including systematic reviews, have still applied the Cox proportional hazard model ^10,11^. It should be evaluated by methods that can be adapted when the assumption is not valid, such as restricted mean survival time (RMST) analysis ^12^.

Kaplan-Meier (KM) survival curves shown in published papers can be considered as easily accessible, highly informative, and useful research materials. We previously analyzed KM curves from the ICON7 ^8^, GOG-0218 ^9^, BOOST ^13^ trials and reported the changes in relative risk of progression between the treatment arms at every 15 months ^14^. In recent years, several methods for more detailed image-based analysis of published KM curves and reconstruction of the original data have been reported ^15, 16^.

Here we analyze patient gene expression data from ancillary analyses of the ICON7 ^17, 18^ (ICON7-A cohort). We also analyze images of KM curves from all the published phase III clinical trials of bevacizumab in both first-line and recurrent ovarian cancer. We then show how the risk of progression changes over time with bevacizumab treatment. These results provide important insights for optimizing the use of bevacizumab in the treatment of ovarian cancer.

## Materials and methods

### ICON7-A cohort

We integrated datasets from two independent ancillary analyses of ICON7, by Kommes et al ^17^ and Desbois et al ^18^. In the dataset by Kommes et al., DASL gene expression microarray analysis was performed on formalin-fixed paraffin-embedded (FFPE) tumor tissues from patients enrolled in the ICON7 trial, and normalized gene expression profiles with clinical information were obtained from the NCBI Gene Expression Omnibus (GEO140082). In the dataset by Desbois et al., total RNA sequencing analysis was performed on archived FFPE tumor tissues and raw sequencing data with clinical information were obtained through the European Genome-Phenome Archive (accession number EGAS00001003487). The downloaded paired-end fastq files were preprocessed using fastp ^19^, mapped on to the human reference genome (GRCh38) using STAR-RSEM ^20,21^, and gene expression values were quantified as fragments per kilobase of exon per million reads mapped (FPKM). Finally, the above gene expression and clinical data were integrated into 745 cases (Supplementary table S1).

### HRDness prediction

We previously identified a gene expression signature associated with HRD (HRDness signature) in ovarian cancer using the dataset from The Cancer Genome Atlas (TCGA-OV), developed a scoring method to calculate its enrichment for each sample and a machine learning method to predict whether each sample is HRD or non-HRD, using an external gene expression dataset as input ^6^. Briefly, TCGA-OV samples were labeled as either HRD or non-HRD based on their genomic scar scores, and the gene expression values of the HRDness signature were used as feature values to build four classifiers using different machine learning methods, namely, k-nearest neighbor, support vector machine, random forest, and linear regression. Input gene expression values for HRDness signature were calibrated against the TCGA-OV dataset using SVA::Combat^22^. In this study, each tumor was assigned to HRD when two or more of the four classifiers predicted HRD; otherwise to non-HRD.

### Validation of HRDness prediction in external FFPE ovarian tumor samples

Raw sequencing data of tumor RNA sequencing and normal-tumor paired whole exome sequencing from 61 FFPE specimens analyzed by Kang et al. ^23^ were obtained from the NCBI Sequence Read Archive (PRJNA700673). Somatic mutation call was performed with Mutect2 using hg38 as the reference genome, referring to the Broad Institute’s GATK4 pipeline: Somatic short variant discovery (SNVs + Indels). For the *BRCA1/2* genes, truncating mutations that were not marked as ‘benign’ or ‘VUS’ in ClinVar ^24^ or InterVar ^25^ were retained. Germline *BRCA1/2* variant call with HaplotypeCaller referring to the Broad Institute’s GATK4 pipeline: Germline short variant discovery (SNPs + Indels). Truncating mutations that passed the hard-filtration and were annotated as ‘pathogenic’ in ClinVar ^24^ or InterVar ^25^ were extracted.

Using the whole exome data as input, Sequenza ^26^ and scarHRD ^27^ were used to calculate the HRD score as the sum of telomeric allelic imbalance (TAI), large-scale state transition (LST), and genomic loss of heterozygosity (LOH) scores.

According to the method in the original paper ^23^, the Signature 3 ratio was calculated from the contribution rate of Signature 3 decomposed by MutationalPatterns ^28^ using all detected somatic mutations as input and COSMIC version 2 mutational signatures 1, 3, and 5 as reference.

### Calculation of progression risk over time using images of published Kaplan-Meier curves

The coordinates of the XY axis and each KM curve were extracted from the figures of the papers using ImageJ software ^29^. Missing parts of the curves were manually completed. The survival rate at each day point on the curve was calculated based on the X-axis coordinates at time 0 and 12 months (or 24 months) and the Y-axis coordinates at survival rates of 0 and 100%. The progression risk at a given time point was calculated as the decrease on the survival curve at 30 days after that time point. The ratio of the progression risk of the subject group to that of the control group at each time point was calculated as the relative risk. When either was zero, the relative risk was considered to be incalculable. The progression risks and relative risk at each time point were smoothed by simple moving averages over the 60 days before and after the time point, and the trends were analyzed.

### Statistical analysis

Restricted mean survival time analysis (RMST) was performed using survRM2 (https://github.com/cran/survRM2). Adjusted RMST by integrating an adjusted Kaplan-Meier estimator with inverse probability weighting was performed as previously reported ^30^. All other statistical analyses and result visualization were performed using Python (3.8.8). Survival analyses including Kaplan–Meier curve, Cox proportional hazard assumption test, and log-rank test were performed using Lifelines (0.26.3). Spearman’s rank correlation test was performed using SciPy (1.7.2). Machine learning analyses were performed using Scikit-learn (1.0.1). A P value <0.05 was considered statistically significant.

### Data availability

Integrated data from the two ancillary studies of ICON7 (ICON7-A) were provided in supplementary table S1. The sources of the KM curves used in the study were summarized in supplementary table S2. Survival rate and progression risk data for each day point calculated from KM curves were summarized in supplementary table S3.

## Results

### Progression risk over time in ICON7-A cohort

In the ICON7-A cohort (n=745) (see methods), comparative analysis of progression-free survival (PFS) between the bevacizumab group (Bev+) and standard treatment group (Bev−) revealed that the proportional hazards model was not valid. (Figure 1A, 1B). We calculated the risk of progression at a given point in time based on the number of patients who progressed during the following 30 days and the ratio of the risk between the two groups (Bev+/Bev−) was calculated as the relative risk (Figure 1C). The trends of these values were examined by smoothing with a simple moving averages from before and after the 60 days (Figure 1D,1E). Interestingly, the risk of progression in the Bev+ group was lower than the Bev-group in the early treatment period, but gradually increased from around 6 months, reached as the same level as the Bev-group at around 12 months, when treatment was discontinued, and exceeded the Bev-group thereafter, showing “rebound effect” (Figure 1F). Restricted mean survival time (RMST) analysis showed that PFS was not significantly different between the two groups in the overall period, but was significantly better in the Bev+ group before bevacizumab discontinuation, and was significantly worse in the Bev+ group after the discontinuation (Figure 1G). In addition, adjusted RMST (ARMST) analysis ^30^ using stage, surgical completion, age, and histology as covariates showed similar results (Figure 2).

**Figure 1.**
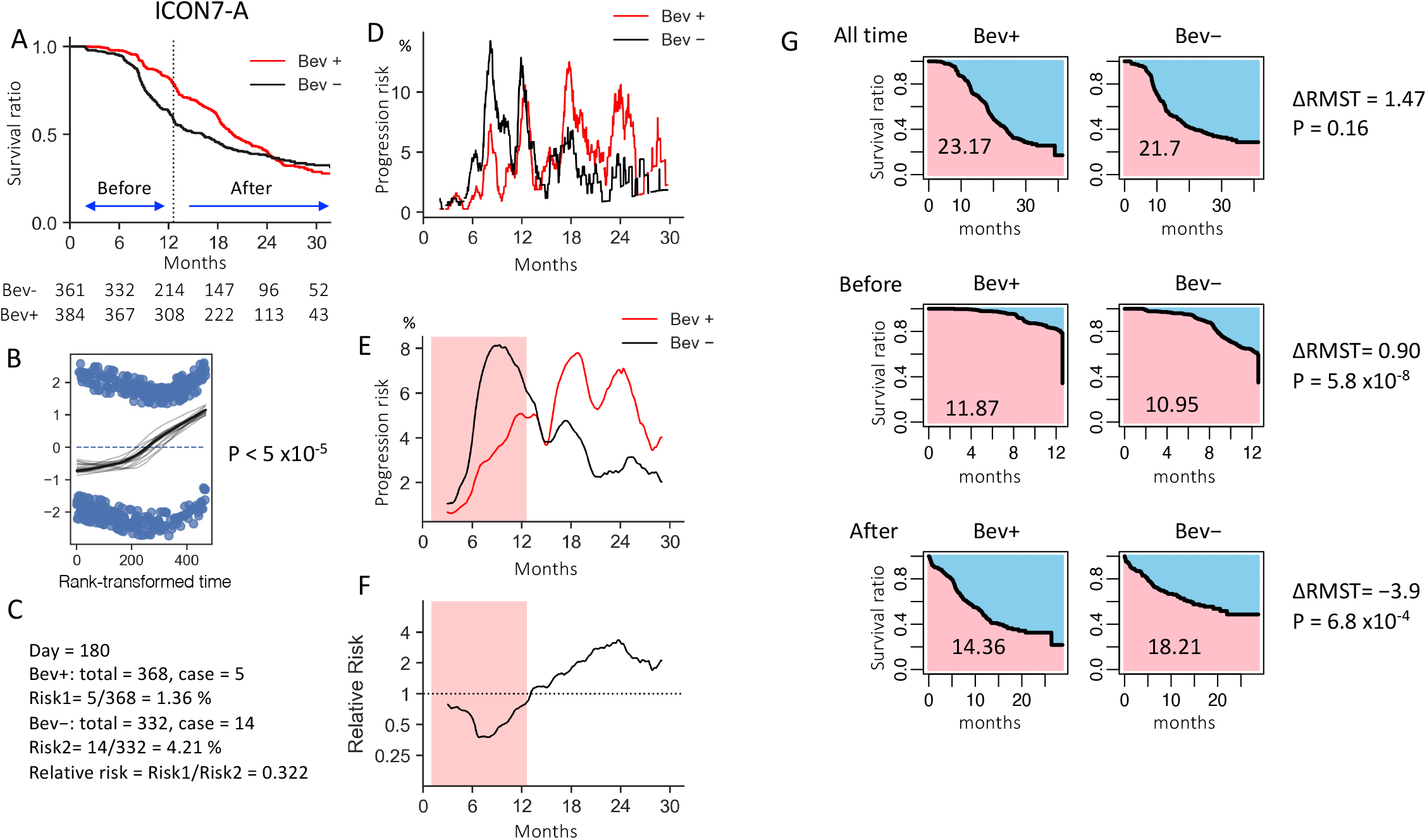
Analysis of the ICON7-A cohort. A) Kaplan-Meier (KM) survival curves stratified by bevacizumab treatment. The dotted line represents the time of bevacizumab discontinuation. The observation period was divided before and after bevacizumab discontinuation. Bev+: bevacizumab treatment group, Bev−: control group B) Test of proportional hazard assumption in the Cox model by the Schoenfeld residuals plot. The hazard ratio between the two groups stratified by bevacizumab treatment is not consistent over time. C) Calculation of the risk of progression at a given point in time: example at day 180 (month 6) from the start of observation. Based on individual patient clinical data, 368 and 332 patients in Bev+ and Bev-were progression-free at day 180, respectively. From that point to 30 days later, 5 and 14 patients had a relapse. Cases that were censored without recurrence were excluded in both groups. The risk of progression was estimated to be 5/368 (=1.36%) in Bev+ and 14/328 (=4.27%) in Bev− and the relative risk was (5/368)/(14/332) = 0.322. D) Plot of the risk of progression per day in the two groups. E) Moving averages of the risk of progression in the two groups. Simple moving averages at 60 days before and after were used at each time point. The red background color represents the period of bevacizumab administration. F) Moving averages of the relative risk of progression between the two groups. G) Comparison of PFS in the two groups by restricted mean survival time (RMST) analysis for the overall period (top) and before (middle) and after (bottom) discontinuation of bevacizumab. PFS was not significantly different between the two groups in the overall period, but significantly longer in Bev+ before bevacizumab discontinuation, but significantly shorter in Bev+ group after the discontinuation.

**Figure 2.**
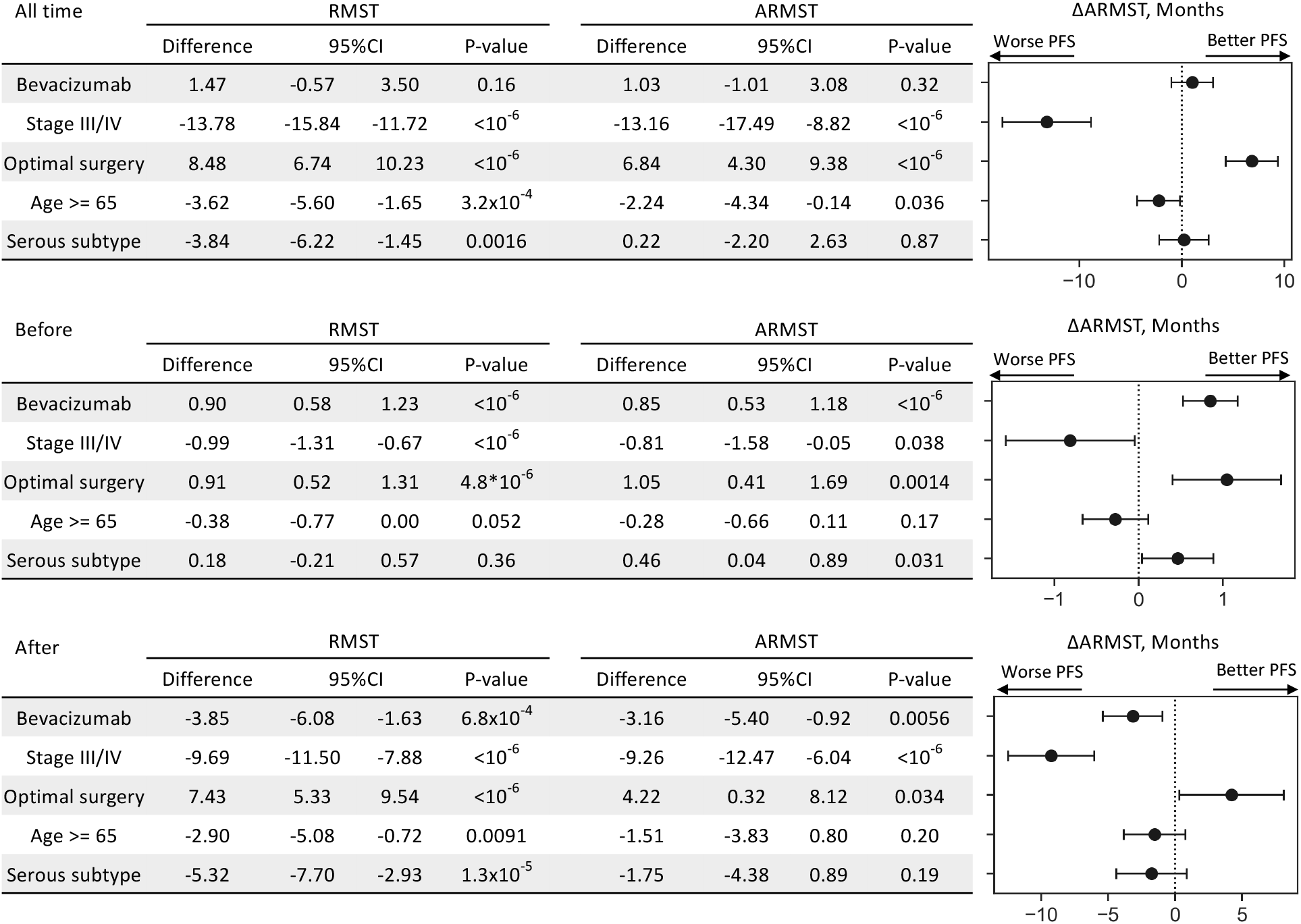
RMST and adjusted RMST (ARMST) analyses of the ICON7-A cohort. Top; the overall period, middle; before bevacizumab discontinuation, bottom; after bevacizumab discontinuation. PFS of Bev+ was not significantly different from Bev-in the overall period but was significantly better than Bev-before bevacizumab discontinuation and worse than Bev-after bevacizumab discontinuation.

### Analysis of ICON7-A divided into serous and non-serous tumors

As shown in Figure2, the serous subtype showed a better PFS before bevacizumab discontinuation but a worse PFS after the discontinuation than the non-serous subtype. We stratified the patients by serous (n=535) and non-serous (n=210) subtypes and compared Bev+ and Bev− groups. In the serous subtype, a similar rebound effect was observed as in the whole cohort (Figure3A,3B,3C) ; the risk of progression in the Bev+ and Bev-groups was reversed before and after the bevacizumab discontinuation (Figure3D). In contrast, in the non-serous subtype, there was a gradual decreased in the risk of progression during bevacizumab treatment period in the Bev+ group but no obvious rebound effect was observed after the discontinuation (Figures 3E-3H).

**Figure 3.**
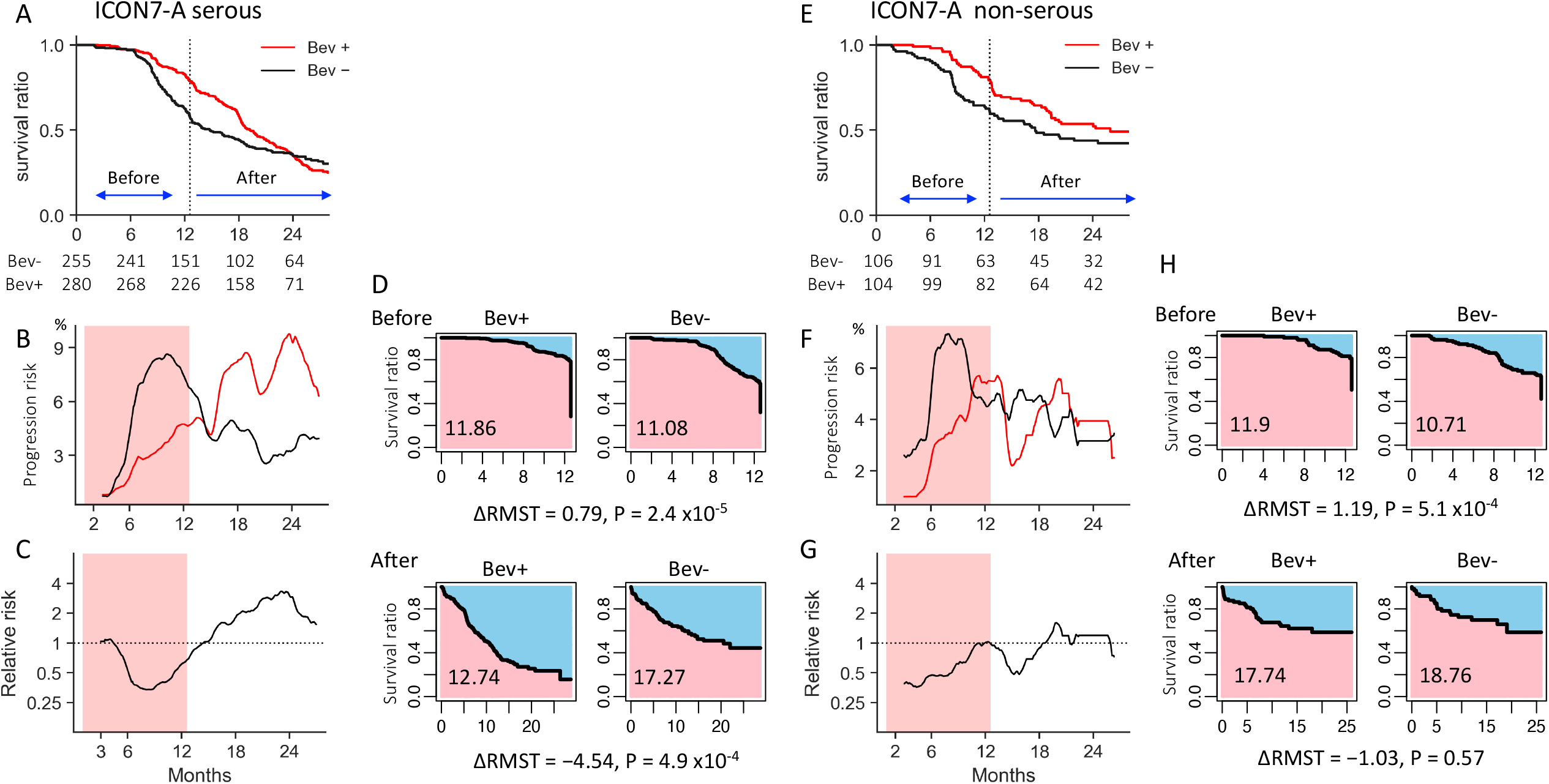
Analysis of the ICON7-A divided into serous and non-serous histology. A) KM survival curves, B) risk of progression, C) relative risk of Bev+ to Bev−, and D) RMST analysis in the serous subtype. Time-dependent changes in the risk of progression and the rebound effect in Bev + were similar to the overall cohort (Figure1). The red background color in B and C represents the period of bevacizumab administration. E) KM survival curves, F) risk of progression, G) relative risk of Bev+ to Bev−, and H) RMST analysis in the non-serous subtype. There was no obvious rebound effect after the discontinuation.

### Analysis of ICON7-A serous tumors divided into HRD and non-HRD

We further examined whether the HRD status is related to the bevacizumab effect in the serous type. We first tested whether our previously reported HRDness signature ^6^ could be applicable to gene expression data derived from FFPE specimens. Analysis of whole exome sequencing and RNA sequencing data (n=61) from FFPE samples of previously reported ovarian serous carcinoma ^23^ (see methods) showed that the HRDness signature enrichment score was positively correlated with the HRD score and Signature 3 ratio (r=0.29, 0.39, p=0.021, 0.0019, respectively, Figure SX1A). A study of 25 patients who received olaparib for platinum-sensitive recurrence showed that PFS after olaparib initiation was better in patients with HRD, including those with BRCA1/2 mutations and/or those predicted to have HRD based on gene expression, than in those without (p=0.0146, Figure SX1B), consistent with our previous results ^6^.

Thus, we predicted HRD status of patients in the ICON7-A cohort based on their gene expression (see methods), and found that approximately half of the patients (279/534) were assigned to the HRD group and PFS was better in the HRD group than in the non-HRD group (p=0.039, Figure SX1C). Stratified analysis showed that the change in risk of progression over time and the rebound effect associated with bevacizumab treatment were similarly observed both in HRD and in non-HRD cases (Figure 4), indicating that HRD status did not impact on the effect of bevacizumab administration.

**Figure 4.**
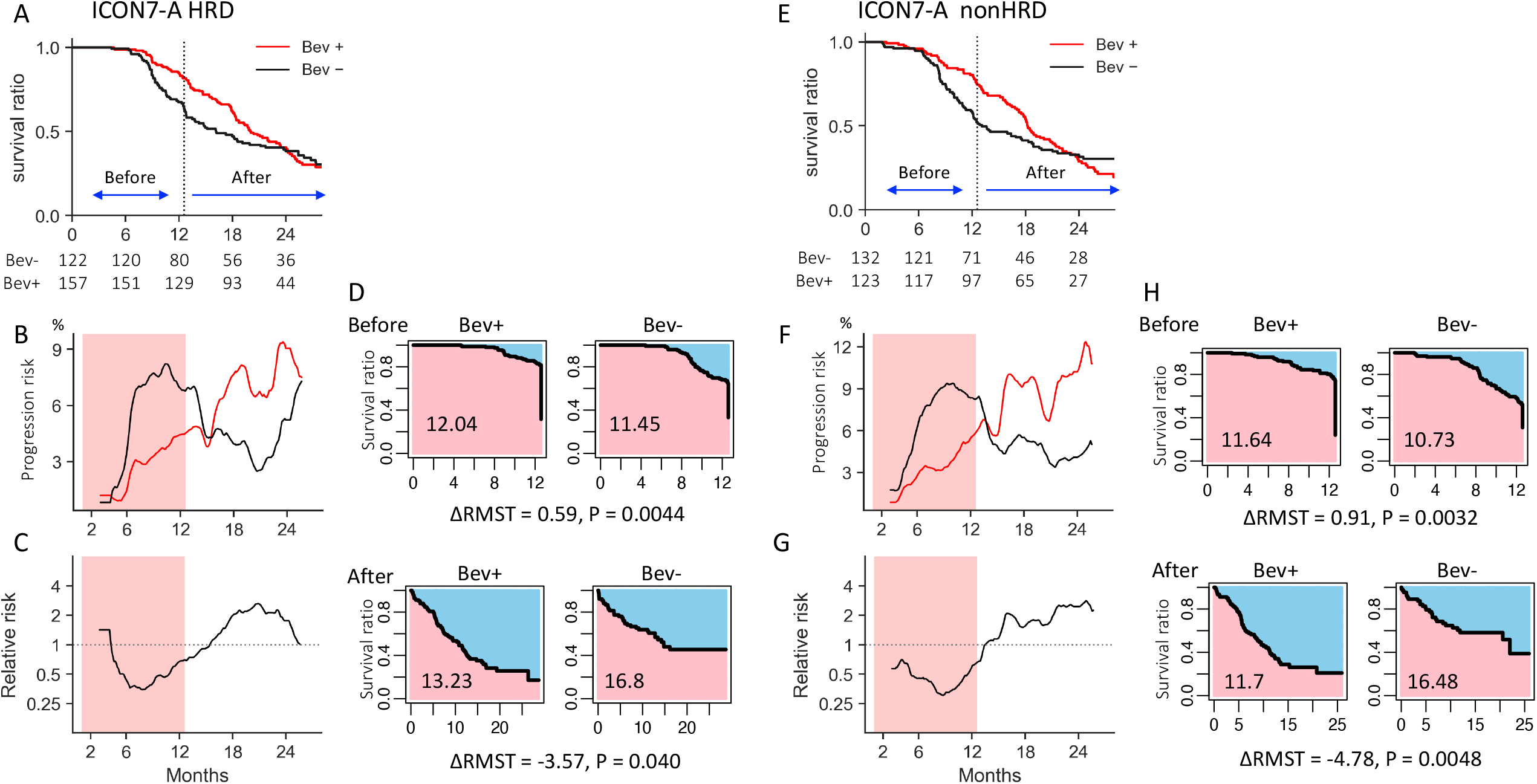
Analysis of the ICON7-A serous subtype divided into HRD and non-HRD tumors. A) KM survival curves, B) risk of progression, C) relative risk of Bev+ to Bev-, and D) RMST analysis in the HRD tumors. E) KM survival curves, F) risk of progression, G) relative risk of Bev+ to Bev-, and RMST analysis in the non-HRD tumors. Time-dependent changes in the risk of progression and the rebound effect in Bev+ were similar to the overall cohort of the serous subtype (Figure3A-3D). The red background color in B, C, F, and G represents the period of bevacizumab administration.

### Analysis of KM curves of first-line treatment cases

Next, we developed a method to estimate event risk at each time point by analyzing images of published Kaplan-Meier (KM) survival curves (see methods, supplementary figure X2A,X2B). The method was applied to the KM curves constructed from individual patient data of the ICON7-A cohort and confirmed to produce a very similar result to the one described above (Figure1DEF, Figure X2CDE).

Using this method, we analyzed the images of the KM curves from the previous phase III trials for bevacizumab (Supplementary table S2). The results from the original ICON7 cohort ^8^ were exactly the same as those from the ICON7-A (Figure5A). The trend was similarly observed both in the high risk cases, defined as those with FIGO stage IV disease or with FIGO stage III disease and >1.0 cm of residual disease after debulking surgery, and in the non-high risk cases (Figure5B). In the BOOST trial ^13^, which randomly assigned patients to receive bevacizumab for either 15 or 30 months (Bev15 or Bev30), the risk of progression in the Bev30 group was slightly lower than in the Bev15 group from month 15 to month 30, but was higher after month 30 (Figure 5C). The results from the GOG-0218 ^9^ (Figure 5D) were similar to those in the ICON7. In a subgroup analysis of GOG-0218 ^31^, in which cases were divided by mutation status in homologous recombination repair-related genes, there was no significant difference in the trends (Figure 5E). The results were also similar in another subgroup analysis of GOG-0218 stratified by chemosensitivity status as determined by changes in blood CA125 levels ^32^ (Figure 5F).

**Figure 5.**
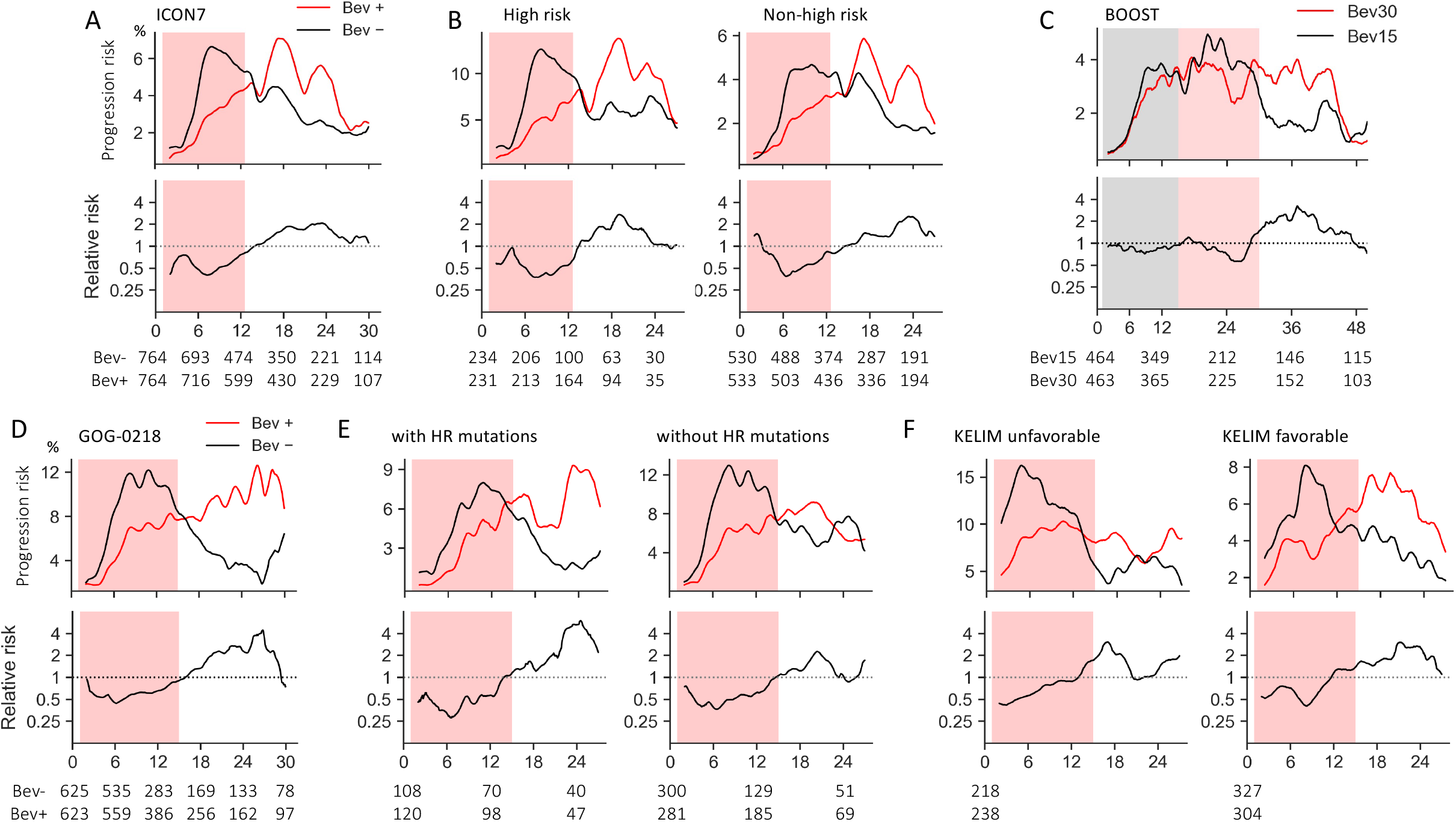
Analyses of KM curves for first-line cases. A) Comparison of the bevacizumab (Bev+) and the control (Bev-) groups in the ICON7 overall cohort. The results were exactly the same as those from the ICON7-A cohort (Figure 2E,2F). The red background color represents the period of bevacizumab administration. B) The high risk (left) and non-high-risk (right) cases in the ICON7 cohort. C) Comparison of bevacizumab for 30 months (Bev 30) and 15 months (Bev 15) in the BOOST cohort. The grey background represents 15 months and red represents 15–30 months of bevacizumab administration. D) GOG–0218 overall cohort. E) GOG–0218 cases with (left) and without (right) homologous recombination repair-related gene mutations. F) GOG–0218 cases with low (left) and high (right) chemotherapy sensitivity based on serum CA125. Changes over time in the risk of progression (upper) and the relative risk (lower) are shown. Time-dependent change in the risk of progression and the rebound effect after discontinuation of bevacizumab were similarly observed in all the above cohorts.

### Analysis of KM curves of recurrent cases

Finally, we analyzed KM curves from the phase III trials in patients with recurrent ovarian cancer (Supplementary table S2). In GOG-0213, bevacizumab was administered in combination with TC followed by maintenance therapy for platinum-sensitive recurrence ^33^. In OCEANS, bevacizumab was administered in combination with gemcitabine plus carboplatin followed by maintenance therapy for platinum-sensitive recurrence ^34^. In AURELIA, bevacizumab was used in combination with a nonplatinum monotherapy for platinum-resistant recurrence ^35^. In MITO16B, bevacizumab was administered in combination with platinum-doublet for platinum-sensitive recurrence previously treated with bevacizumab in the first-line setting ^36^. In all of these studies, the duration of bevacizumab administration was not predetermined and was continued until disease progression or an unacceptable adverse event occurred. In common with all these studies, the relative progression risk of the Bev+ group compared to the Bev-group was lowest soon after the start of treatment and then gradually increased over time, but did not consistently exceed 1, indicating no rebound effect (Figures 6A-D).

**Figure 6.**
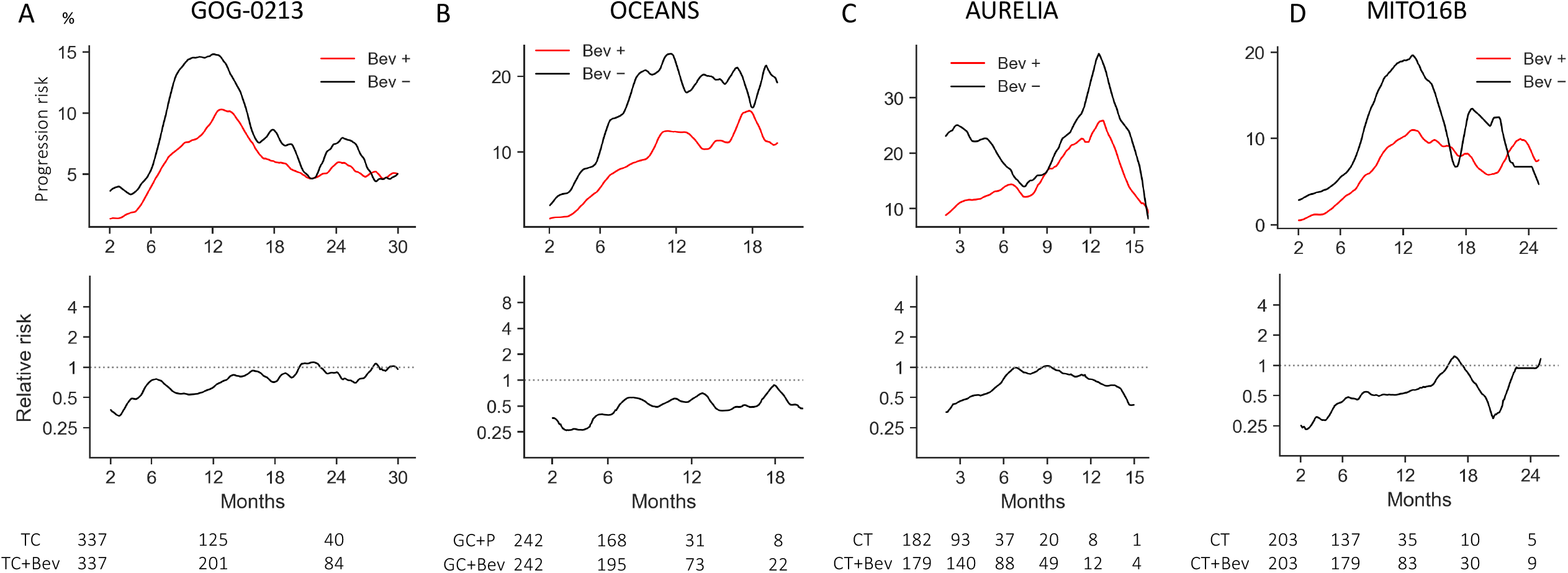
Analyses of KM curves for recurrent cases. A) GOG-0213. B) OCEANS. C) AURELIA. D) MITO16B. Changes over time in the risk of progression (upper) and the relative risk (lower) of the bevacizumab (Bev+) to the control (Bev−) groups are shown. In the recurrent cases, bevacizumab was continued until progression, and no rebound effect was observed.

## Discussion

The ICON7-A cohort (n=745) we compiled in this study had about half the number of cases of the original ICON7 cohort (n=1528) ^8^, and the Kaplan-Meier (KM) curves for PFS was almost identical to that of the original paper (Figure 1A). The effect of bevacizumab on reducing the risk of progression peaked at about 6 months and then disappeared at about 12 months. Even for cancer types other than ovarian cancer, the difference in PFS with bevacizumab is often greatest around 12 months ^2^. The main action of bevacizumab, an anti-VEGF-A antibody, is inhibition of angiogenesis in tumor tissue. Theoretically, it can make tumor tissue hypoxic and hypo-nutrient and induce apoptosis and necrosis of tumor cells, but it has little or no direct cell killing effect ^37^. Instead, the hypoxia-induced, VEGF-independent, delayed angiogenesis that would occur during bevacizumab treatment may be responsible for tumor progression and recurrence ^37^. Early studies reported that restoration of vessel structure and function by anti-VEGF antibodies may improve blood perfusion and drug delivery of cytotoxic agents to tumors ^38^, but more recent studies reported that the combination of anti-VEGF antibodies rather reduced the intratumor concentrations of cytotoxic agents ^39^. And the combination with bevacizumab in the GOG-0218 study did not show any improvement in response rate ^40^. One study reporting that VEGF had tumor immunosuppressive effects ^41^ led to the expectation that anti-VEGF antibodies would activate anti-tumor immunity, but another study reported that hypoxia induced by anti-VEGF antibodies rather suppressed anti-tumor immunity ^42^. Collectively, current evidence suggests that the primary action of bevacizumab is presumed to be solely cytostatic, rather than cytotoxic.

In this study, the ICON7-A cohort analysis stratified by serous and non-serous subtypes showed that the rebound effect was only observed in the serous type but not in the non-serous type (Figure 3). To the best of our knowledge, this is the first report to show differences in the efficacy of bevacizumab between histological subtypes of ovarian cancer. Bevacizumab is thought to be most effective when tumor cell growth is directly dependent on VEGF ^37^, and in some cases of the serous type, cancer cells have been reported to express high levels of VEGF receptors ^43, 44^. In other words, differences in dependence on VEGF among histological subtypes may be related to the efficacy of bevacizumab. In addition, the serous type often responds well to initial chemotherapy but more frequently relapses with increased treatment resistance afterword than the non-serous subtype ^45^. This clinical characteristic of the serous type may also be relevant to the differences in results from the other histological types.

The results of the subgroup analyses of ICON7 and GOG0218 suggested that the change in the progression risk ratio over time and the rebound effect were independent of whether the patient was at high or low risk, with or without HRD, and sensitive or resistant to chemotherapy. Given that bevacizumab is effective for only about a year and has a rebound effect after its discontinuation, it is likely that the benefit of bevacizumab, including improved survival and quality of life, will only be seen in patients with short survival. Our findings may explain the results of previous studies that bevacizumab did not prolong overall survival (OS) in the ICON7 and GOG-0218 overall cohorts ^46, 47^ but did prolong OS in chemotherapy-refractory, high-risk patients ^32, 48^.

The absence of rebound effect in the studies for recurrent ovarian cancer (Figure 6A, B, C) seems to be attributed to the protocol that did not stop bevacizumab until disease progression ^34, 33, 35^. And a similar result was observed in the MITO-16B trial ^36^ where bevacizumab was given again in recurrent disease (Figure 6D). This suggests that bevacizumab is a growth inhibitory agent and does not induce clonal selection in recurrent tumors like cytotoxic agents (^2^). Given that patients who relapse have shorter median PFS and OS than those on first-line treatment, bevacizumab may provide more benefit to patients with recurrent disease.

In the PAOLA-1 trial, in combination with bevacizumab, olaparib significantly prolonged PFS in patients with HRD compared with placebo ^49^. And in patients with no residual tumor after primary debulking surgery, the 2-year PFS in the olaparib group was remarkably favorable: 96% in *BRCA* mutated cases and 80% in HRD cases with wild-type *BRCA* ^50^. A Phase III trial is currently underway to evaluate the efficacy of adding bevacizumab in the presence of niraparib (NCT05009082). The results of this study will clarify whether PARP inhibitor maintenance therapy can reduce the progression after bevacizumab discontinuation.

A limitation of this study is that individual patient data for the entire cohorts of the ICON7 and GOG-0218 trials were not available. This may lead to potential bias, especially in the subgroup analysis with small numbers of cases, such as the non-serous type. In addition, the image analysis of the KM curve used in this study did not allow for statistical analysis. The results of this study need to be verified in future studies.

In conclusion, we have shown that the efficacy of bevacizumab administration on the risk of ovarian cancer progression varies over time. Considering that the rebound effect occurs after completion of bevacizumab in the first-line treatment of serous ovarian cancer, bevacizumab should be most effective in patients who are less likely to be affected by the rebound effect, i.e., those with an expected survival of less than one year. The effect of bevacizumab in combination with PARP inhibitors needs further investigation.

## Data Availability

All data produced in the present work are contained in the manuscript and supplementary data.

## Authors’ contributions

Conceptualization: NM ; Methodology: ST, NM ; Formal analysis and investigation: ST ; Validation: KY, JH ; Writing–original draft preparation: ST, NM; Writing–review and editing: ST, HN, NM, KY, JH, MM ; Resources: ST, KY; Data Curation; ST, NM ; Supervision: JH, MM; Funding acquisition: HN

## Acknowledgements

This study was supported by Japan Society for the Promotion of Science (JSPS) KAKENHI grant number 22K09630 (Grant-in-Aid for Scientific Research C for Hidekatsu Nakai). We would like to thank Dr. Hyeon Gu Kang for providing additional information on their published data.

