## Supplementary Figures for "Bevacizumab in ovarian cancer: time-dependent changes in risk of progression"

### Supplementary appendix

#### Supplementary figure X1

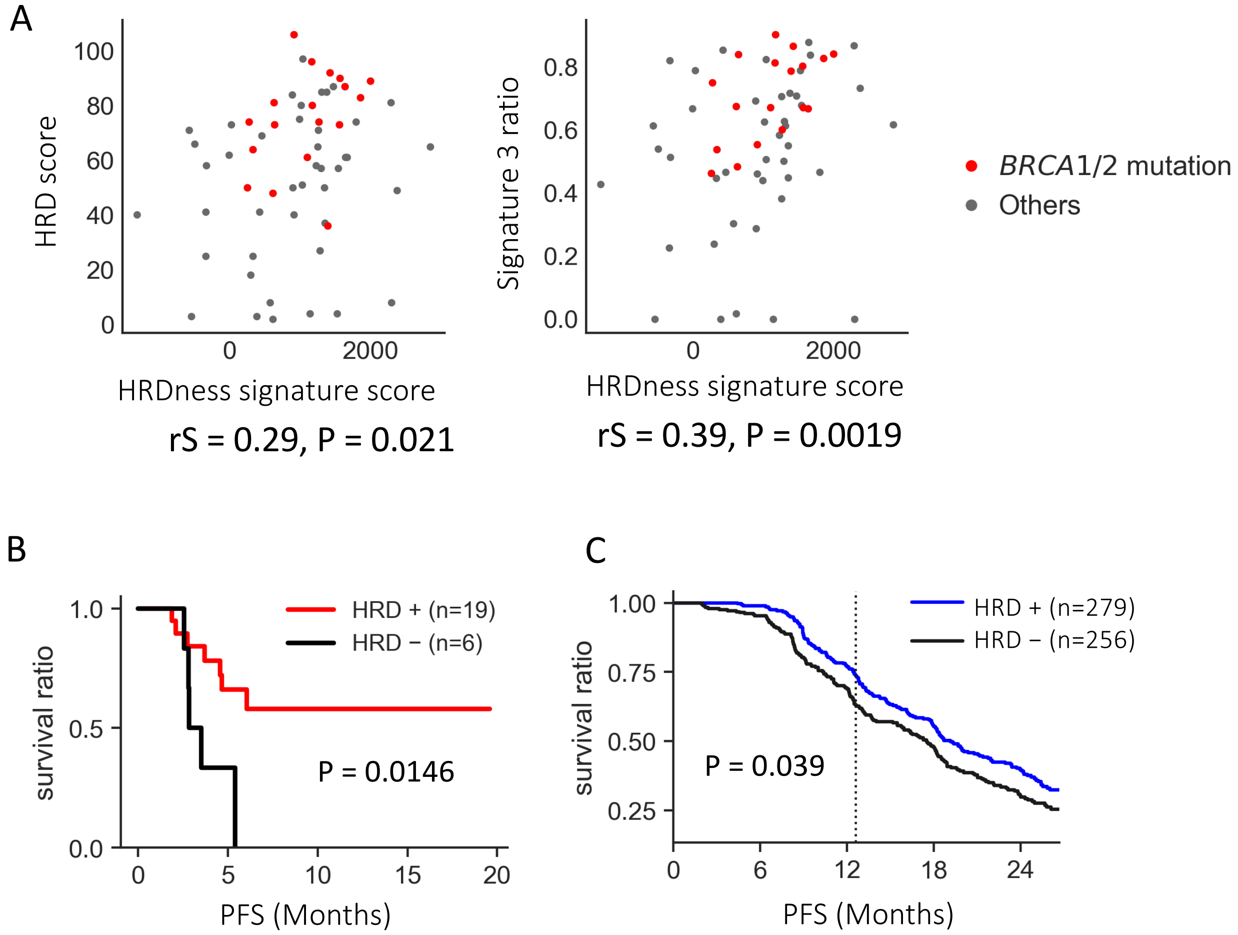


**HRDness prediction in FFPE ovarian tumor samples.**

A) Association between the HRDness signature enrichment score and the genomic scar scores using 61 previously published FFPE-derived serous ovarian cancer samples. (PMID:34711610). The HRDness signature score was positively correlated with the HRD score (left) and Signature 3 ratio (right) calculated from the whole exome sequencing data. rS; Spearman' s correlation coefficient, P; Spearman' s correlation p-value.

B) Survival analysis after olaparib initiation in the patients with platinum-sensitive recurrence. PFS was significantly better in patients with HRD, including those with BRCA1/2 mutations and/or those assigned by the HRDness prediction, than in the rest. P; p-value based on the log-rank test.

C) Survival analysis for PFS between groups of ICON7-A cohort divided by the HRDness prediction. Tumors classified as HRD by the HRDness prediction had a better prognosis than the others. P; p-value based on the log-rank test.

**Supplementary figure X2**

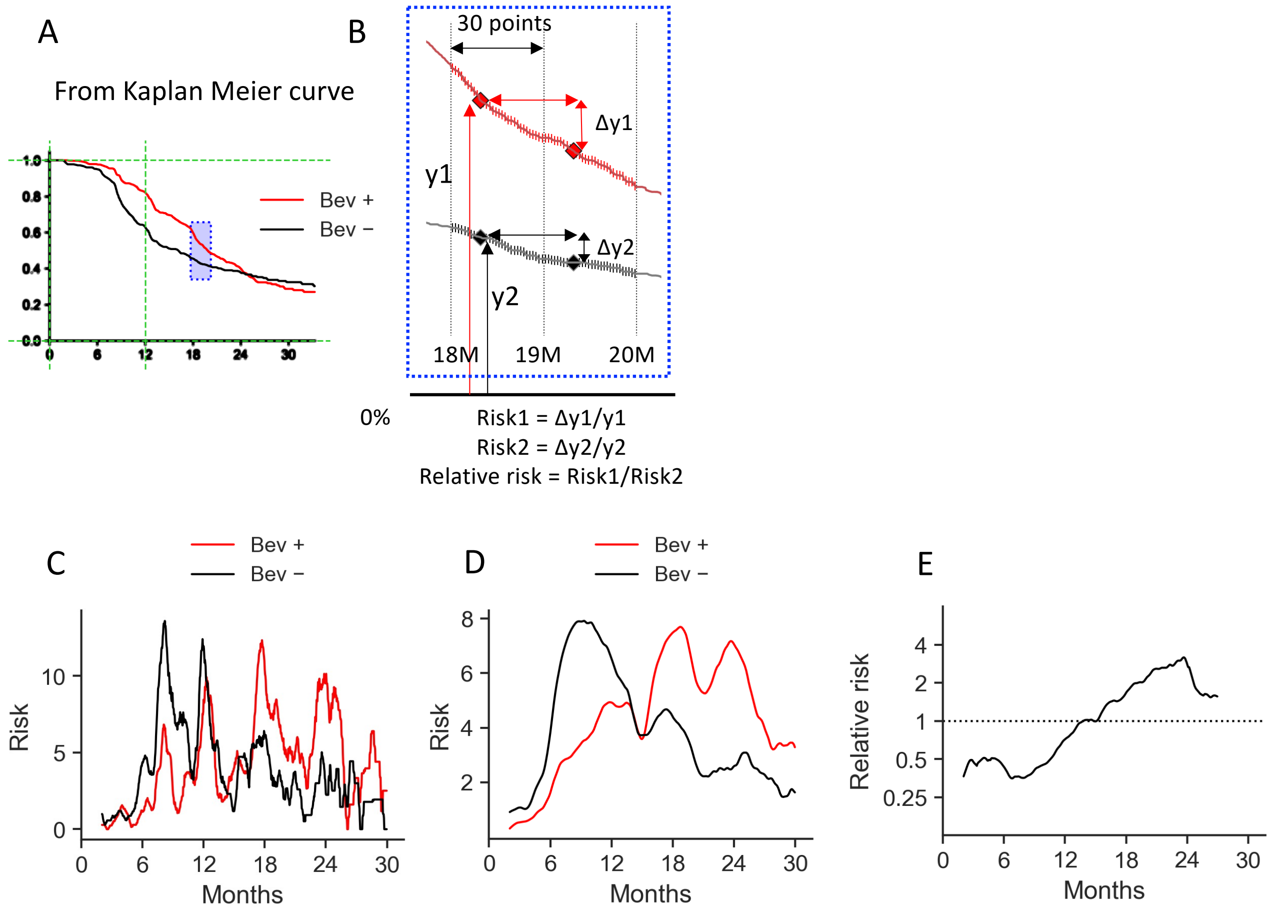


**Validation of the KM curve image-based analysis in the ICON7-A cohort.**

A) Reconstruction of KM curves using Image J.
The survival rates at each day point on the KM curves of the bevacizumab (Bev+) and the control group (Bev-) were calculated based on the X-axis coordinates at time 0 and 12 months and the Y-axis coordinates at survival rates of 0 and 100%.

B) Calculation of the risk of progression at a given point in time: example at one day after 18 months (enlarged view of the area with blue background in A).
The progression risk at a given time point was calculated as the decrease on the survival curve at 30 days after that time point. The relative risk was calculated as the ratio of the progression risk of the subject group to the control group at each time point.

C) Plot of the risk of progression per day in the two groups.

D) Moving averages of the risk of progression in the two groups.
Simple moving averages at 60 days before and after were used at each time point.

E) Moving averages of the relative risk of progression between the two groups.
